# Characterising personal, household, and community PM2.5 exposure in one urban and two rural communities in China

**DOI:** 10.1101/2023.04.10.23288228

**Authors:** Ka Hung Chan, Xi Xia, Cong Liu, Haidong Kan, Aiden Doherty, Steve Hung Lam Yim, Neil Wright, Christiana Kartsonaki, Xiaoming Yang, Rebecca Stevens, Xiaoyu Chang, Dianjianyi Sun, Canqing Yu, Jun Lv, Liming Li, Kin-Fai Ho, Kin Bong Hubert Lam, Zhengming Chen, China Kadoorie Biobank collaborative group

## Abstract

**Background:** Cooking and heating in households contribute importantly to air pollution exposure worldwide. However, there is insufficient investigation of measured fine particulate matter (PM_2.5_) exposure levels, variability, seasonality, and inter-spatial dynamics associated with these behaviours.

**Methods:** We undertook parallel measurements of personal, household (kitchen and living room), and community PM_2.5_ in summer (May-September 2017) and winter (November 2017-Janauary 2018) in ∼480 participants from one urban and two rural communities in China. These recorded ∼61,000-81,000 person-hours of processed data per microenvironment. Age- and sex-adjusted geometric means of PM_2.5_ were calculated by key participant characteristics, overall and by season. Spearman correlation coefficients between PM_2.5_ levels across different microenvironments were computed.

**Findings:** Overall, 25.1% reported use of solid fuel for both cooking and heating. Solid fuel users had ∼90% higher personal and kitchen 24-hour average PM_2.5_ exposure than clean fuel users. Similarly, they also had a greater increase (∼75% vs ∼20%) in personal and household PM_2.5_ from summer to winter, whereas community levels of PM_2.5_ were 2-3 times higher in winter regardless of fuel use. Compared with clean fuel users, solid fuel users had markedly higher weighted annual average PM_2.5_ exposure at personal (77.8 [95% CI 71.1-85.2] vs ∼40 µg/m_3_), kitchen (103.7 [91.5-117.6] vs ∼50 µg/m_3_) and living room (62.0 [57.1-67.4] vs ∼40 µg/m_3_) microenvironments. There was a remarkable diurnal variability in PM_2.5_ exposure among the participants, with 5-minute moving average 700-1,200µg/m^3^ in typical meal times. Personal PM_2.5_ was moderately correlated with living room (Spearman r: 0.64-0.66) and kitchen (0.52-0.59) levels, but only weakly correlated with community levels, especially in summer (0.15-0.34) and among solid fuel users (0.11-0.31).

**Conclusion:** Solid fuel use for cooking and heating was associated with substantially higher personal and household PM_2.5_ exposure than clean fuel users. Household PM_2.5_ appeared a better proxy of personal exposure than community PM_2.5_ in this setting.

## 1. Introduction

The growing population and energy demand from rapid urbanisation, coupled with continued reliance of fossil fuels, have aggravated the ambient air pollution in many low-and middle-income countries (LMICs). On the other hand, about 3 billion individuals are still relying on solid fuels (e.g. coal, wood) for cooking and heating, which can result in intensive household air pollution.^1, 2^ Fine particulate matter (PM_2.5_) from domestic use of solid fuels and ambient sources together constitute the top environmental risk factor of disease burden globally, estimated to account for more than 6 million premature deaths in 2019._1_ Despite the global health significance,_3,4_ there remains substantial uncertainties in the exposure-disease relationships and thus disease burden estimation, as most existing epidemiological studies relied on exposure proxies, namely modelled ambient air pollution levels around residential addresses and self-reported fuel use for indoor or household air pollution exposure._3,5_

Until recently, directly measured air pollution exposure data are rarely available in large population-based epidemiological studies. Most measurement studies had relatively small sample sizes, assessed primarily kitchen PM_2.5_ levels, and were limited to one or a few rural communities, with limited repeated measurements across seasons._6-14_ The largest relevant study to date (PURE-Air) collected 48-hour aggregated kitchen and personal PM_2.5_ data in ∼2400 households and ∼900 individuals, respectively, in rural areas from eight LMICs.^6^ They found substantial variability in kitchen and personal PM_2.5_ levels by cooking fuel types and across countries, with solid fuel users tend to show significantly higher exposure. However, the short measurement window, limited repeated seasonal measurements, and inadequate coverage of heating season exposure leave ambiguity to both within-week and seasonal exposure variability within and between individuals. There is also a need of time-resolved data and parallel assessment of not only personal and kitchen PM_2.5_ but also living room and ambient levels to better understand the spatial-temporal dynamics of PM_2.5_ exposure. Data from urban areas will also offer additional insight into the urban-rural contrast in PM_2.5_ exposure patterns.

We report detailed analyses of questionnaire data on personal characteristics, fuel use, and time-resolved PM_2.5_ exposure data at personal, household, and community levels in ∼480 participants from one urban and two rural areas in the China Kadoorie Biobank (CKB), repeatedly in the warm and cool seasons._15_ The present report aims to i) examine both aggregated and time-resolved PM_2.5_ levels by fuel use and other key characteristics; and ii) clarify personal-household-community gradient of PM_2.5_ exposure.

## 2. Materials and Methods

### 2.1 Study design and sample

CKB is an ongoing prospective cohort study of ∼512,000 adults aged 30-79 years recruited from ten diverse areas of China during 2004-2008._16,17_ The CKB-Air study was nested within CKB, and details of the design, data collection procedures, data cleaning and processing, and participant characteristics have been published previously._15_ Briefly, 488 participants (mean age 58 years, 72% women) were recruited from two rural (Gansu, Henan) and one urban (Suzhou) CKB study sites (**eFigure 1**), selected to capture a diverse range of fuel use patterns._18_ The study involved repeated assessment of air pollution and time-activity in the warm (May–September 2017; hereafter referred to as ‘summer’) and cool (November 2017–January 2018; ‘winter’) seasons, with a household questionnaire on participant characteristics and usual fuel use patterns administered in winter. Subsequently, 451 individuals participated in the summer assessment, of whom 37 were not available in winter and were replaced by other eligible CKB participants in the same community. The participants included in the two seasons were similar in their socio-demographics and lifestyle characteristics documented in 2004-2008 during the baseline assessment._15_

The study was approved by the Oxford University Tropical Research Ethics Committee, Oxford, UK (Ref: 5109-17) and the institutional review board of Fuwai Hospital, Chinese Academy of Medical Sciences, Beijing, China (Ref: 2018-1038). All participants provided written informed consent upon recruitment.

### 2.2 Questionnaire data

Trained health workers administered a laptop-based household questionnaire in the cool season, to assess personal characteristics (age, sex, household income, occupation, smoking, environmental tobacco smoke exposure) and exposure to household air pollution (cooking and heating patterns and all fuel types used) (**Text S2**). For those who reported different cooking patterns or fuel use in summer, additional questions on exposure during summer were asked. While many previous studies focused on a single primary cooking or heating fuels, we attempted to capture the increasingly recognised ‘fuel stacking’ phenomenon by assessing all fuel types used._18_ Cooking fuel combinations were derived based on all fuel types reported to be ‘used in most meals’ or ‘sometimes’. A similar approach was undertaken to derive ‘heating fuel combination’ based on the duration of heating fuel use during winter. Clean fuels include gas, electricity, solar, and city-wide district heating (for heating only); solid fuels include coal (smoky/smokeless), coal briquette, charcoal, wood, and crop residue. The electronic questionnaire have built-in error and logic checks to minimise missing data and human errors.

### 2.3 Air pollution data

#### 2.3.1 Air pollution monitors

The study involved ∼120 consecutive hours of measurements (at 1-minute resolution) of fine particulate matter (PM_2.5;_ µg/m^3^) levels, temperature, and relative humidity (%) in three different microenvironments (personal, kitchen, and living room) for each participant, both in summer and winter except for those who only participated in one season (n=74). The measurements were taken using PATS (Particle and Temperature Sensor; Berkeley Air Monitoring Group, CA, USA), an internationally validated low-cost nephelometer-based device (R_2_ range: 0.90-0.99 with reference to both with well-established time-resolved instruments [e.g. TSI DustTrak] and gravimetric measurements) developed for high household air pollution settings (PM_2.5_ detection range: 10-30,000 µg/m^3^).^19^ At each study site, community air pollution (PM_1_, PM_2.5_, PM_10_, carbon monoxide, ozone, and nitrogen oxides) was measured on the roof top of a building in a central location away from any proximal sources of pollution, using two tailor-made research instruments (NAS-AF100; Sapiens Environmental Technology, Hong Kong, China).

Details of quality control and device calibration have been described previously._15_ In brief, all devices were factory-calibrated against wood smoke by the manufacturers, and further calibrated using filter-based personal and static samplers for PM measurements.

Randomly selected PATS (n=15) were also tested for consistency through co-location comparison tests in controlled settings for 24 hours, with good agreement demonstrated (correlation coefficients: 0.85-0.99). Before and after each deployment, the PATS devices were calibrated against HEPA-filtered air for 10 minutes, following the manufacturer’s standardised procedures.

#### 2.3.2 Data cleaning and processing

Participants with corrupted data files due to human or device error were excluded from the PM_2.5_ analyses (**eFigure 2**). Twenty-nine and 115 participants in the warm and cool seasons, respectively, had no community air pollution data from NAS-F100 due to delays in deployment or other logistical challenges (**eFigure 2**). The time-resolved PM_2.5_ data from each PATS were then inspected and processed by i) downsizing to 5-minute moving averages time-series to facilitate computation, ii) applying 20-minute moving median smoothing to replace sporadic extreme spikes, and iii) adjusting data points at persistently high or low (i.e. at the lower limit of detection of 10µg/m_3_) levels by cross-device calibration. Specifically, potentially erroneous data from one device were replaced by imputed data based on levels recorded in the other devices using generalised linear regression. The persistently high levels were likely caused by particles lodged inside the nephelometer or sustained direct light impact; whereas the persistently low levels, most of which were found in the personal PATS data during winter, are likely due to an obstructed air inlet (e.g. covered by clothing) or that the PATS was placed inside an enclosed environment (e.g. in a drawer when participants took it off during bathing or sleep). Overall, only <5% of the PM_2.5_ data recorded were flagged as persistently high or low, indicating generally high data quality (**eTable 1**).

We first removed data from the first and last hour of the measurement period when participants’ exposure was likely affected by the study procedures. We then removed participants with <24 hours of effective data, which could happen due to battery failure. Subsequently, each of the remaining participants had at least 24 hours’ worth of data per PATS per season (median_summer_ [Q1-Q3]: 117 [105-119] hours, median_winter_ [Q1-Q3]: 113 [94-117] hours) (**eTable 2**). To further enhance the quality of the analytical dataset, we undertook a conservative approach to remove participants (n=36) with >50% data flagged as persistently high or low in any one PATS (regardless of the quality of the other two device), thereby restricting the analyses to participants with data of satisfactory quality across all three PATSs (n_summer_=419 [92.7%]; n_winter_=365 [81.1%]). Thirty-five participants in summer who did not provide household questionnaire data were further excluded.

The numbers of participants excluded at each stage of data analysis are shown in **eFigure 2**. After data cleaning, the primary analyses on PM_2.5_ included 384 participants, with a total of 80,980 person-hours of PM_2.5_ data each at the personal, kitchen, and living room, and 67,326 person-hours of data at the community level (**eTable 2**).

### 2.4 Data analysis

Using linear regression, we estimated age- and sex-adjusted (where appropriate) geometric means and 95% confidence intervals (CI) of personal and household (kitchen and living room) PM_2.5_ concentrations levels, by demographic characteristics (age, sex, study area, education, occupation, smoking, household size) and household air pollution-related exposures (cooking frequency, self-reported ‘smoky home’ while cooking or heating, and cooking and heating fuel combinations), stratified by season. We also averaged the time-resolved data to produce season-specific 24-hour PM_2.5_ time-series at personal, household, and community levels, according to different cooking and heating fuel combinations.

We obtained regional temperature data during 2005-2017 (corresponding to the follow-up period of the CKB cohort up till the commencement of CKB-Air) from local meteorological offices and calculated the proportion of months with an average temperature <10°C in each region (0.25 for Suzhou, 0.42 for Gansu, 0.33 for Henan), which was used as a weighting coefficient to approximate heating fuel usage. We then estimated microenvironment-specific annual PM_2.5_ exposure levels as a weighted average of exposure levels across summer and winter, by cooking and heating fuel combinations: *annual average_ij_ = w^ij^ * (1-p_k_) + c_ij_ * p_k_*, where *wij* is the summer average and *cij* the winter average for microenvironment *i* (personal, kitchen, living room, or community) among participants in category *j* of cooking and heating fuel combination (no cooking or heating, clean fuels only, any solid fuels), and *pk* is a region-specific weighting coefficient of heating fuel usage described above.

As a preliminary investigation to understand the relationships between PM_2.5_ levels across microenvironments and by season, we have examined the season-specific Spearman correlation of log-transformed PM_2.5_ levels across the four microenvironments overall and by cooking and heating fuel combinations.

### Role of the funding source

The study funders had no role in study design, data collection, analysis, interpretation, or writing of the report. KHC, XX, KH, KBHL, and ZC had access to all data and had final responsibility for the decision to submit for publication.

## 3. Results

### 3.1 Basic characteristics and PM_2.5_ levels

Of the 384 participants included in the main analyses, the mean age was 58.2 [SD 6.6] years, 74.7% were women, and 32.0% and 52.2% used solid fuels for cooking and heating, respectively. In particular, those who used solid fuels for cooking were more likely to be women, from rural areas, less educated, agricultural workers or home-makers, and to use solid fuel for heating (**eTable 3**). Moreover, substantially more solid fuel users reported observing a smoky home while cooking or heating compared to clean fuel users.

Overall, levels of exposure to PM_2.5_ were generally higher in younger (<65 years) participants, women, and those with lower education, with markedly higher levels in winter than in summer (**Table 1**). Agricultural workers, active smokers, and participants who reported a smoky home while cooking had particularly high PM_2.5_ exposure, most notably at personal and kitchen levels, both in summer and winter. For example, average kitchen PM_2.5_ in summer for those observing smoky home while cooking was 53.7 [95% CI 49.7-58.0] compared to 40.9 [38.5-43.5] for those without such observation, and in winter 119.5 [107.7-132.5] µg/m_3_ compared to 61.8 [56.6-67.6] µg/m_3_. Participants who reported smoky home while heating had somewhat lower personal and living room PM_2.5_ levels in summer, but significantly higher personal (82.0 [74.9-89.7] vs 55.3 [51.2-59.8] µg/m_3_), kitchen (127.9 [114.8-142.4] vs 72.6 [66.2-79.6] µg/m_3_), and living room (72.5 [66.4-79.2] vs 54.2 [50.2-58.4] µg/m_3_) levels in winter. There was no clear pattern by household size.

**Table 1.** Age- and sex-adjusted geometric mean (95% CI) PM2.5 concentrations (μg/m^3^) recorded in the personal, kitchen, and living room monitors by season and key characteristics.

| Characteristics | Summer |  |  |  | Winter |  |  |  |
| --- | --- | --- | --- | --- | --- | --- | --- | --- |
|  | N* | Personal | Kitchen | Living room | N* | Personal | Kitchen | Living room |
| **Personal characteristics** |  |  |  |  |  |  |  |  |
| **Age<sup>†</sup>** |  |  |  |  |  |  |  |  |
| < 65 years | 323 | 42.0 (40.2-43.9) | 43.9 (41.8-46.2) | 33.5 (32.1-34.9) | 305 | 59.5 (56.2-63.1) | 82.8 (77.2-88.8) | 56.1 (52.9-59.4) |
| ≥ 65 years | 61 | 33.5 (30.7-36.5) | 43.0 (39.0-47.5) | 31.9 (29.4-34.6) | 59 | 48.9 (43.6-54.9) | 62.9 (54.7-72.2) | 49.2 (43.9-55.1) |
| **Sex<sup>‡</sup>** |  |  |  |  |  |  |  |  |
| Female | 287 | 40.9 (39.3-42.6) | 44.4 (42.4-46.5) | 32.7 (31.4-33.9) | 267 | 66.1 (62.6-69.8) | 87.6 (82.1-93.5) | 61.1 (57.9-64.5) |
| Male | 97 | 39.8 (37.2-42.7) | 43.3 (40.1-46.9) | 33.7 (31.6-36.0) | 97 | 50.2 (45.8-55.0) | 72.0 (64.5-80.4) | 49.4 (45.1-54.1) |
| **Education** |  |  |  |  |  |  |  |  |
| No formal education | 97 | 48.9 (45.1-53.1) | 52.3 (47.6-57.5) | 38.2 (35.3-41.3) | 92 | 75.8 (67.9-84.7) | 112.6 (98.7-128.5) | 70.2 (62.9-78.3) |
| Primary & middle school | 133 | 38.8 (36.5-41.4) | 42.8 (39.8-46.0) | 30.4 (28.7-32.3) | 125 | 58.5 (53.8-63.5) | 84.0 (76.1-92.8) | 58.4 (53.8-63.4) |
| High school or above | 154 | 38.4 (36.3-40.7) | 41.6 (39.0-44.4) | 33.3 (31.5-35.1) | 147 | 50.9 (47.1-55.0) | 66.0 (60.2-72.4) | 47.4 (44.0-51.2) |
| **Occupation** |  |  |  |  |  |  |  |  |
| Agricultural worker | 140 | 53.2 (50.3-56.3) | 59.9 (56.2-63.9) | 38.0 (36.0-40.2) | 138 | 74.6 (69.2-80.4) | 120.0 (109.6-131.3) | 69.0 (63.9-74.4) |
| Factory worker | 20 | 34.4 (29.7-39.9) | 39.1 (33.0-46.2) | 31.7 (27.4-36.6) | 18 | 59.0 (47.9-72.7) | 59.2 (46.0-76.1) | 36.9 (29.9-45.5) |
| Home-maker | 106 | 40.1 (37.3-43.2) | 43.2 (39.7-47.0) | 32.9 (30.6-35.4) | 109 | 68.6 (62.3-75.5) | 88.4 (78.8-99.2) | 65.3 (59.3-72.0) |
| Non-manual labour | 9 | 33.4 (27.0-41.4) | 29.0 (22.7-37.0) | 26.2 (21.2-32.4) | 10 | 62.0 (47.1-81.8) | 46.0 (33.0-64.2) | 45.2 (34.2-59.7) |
| Self/ un-employed or other | 109 | 29.1 (27.2-31.1) | 30.7 (28.4-33.1) | 28.5 (26.7-30.5) | 89 | 31.6 (28.7-34.8) | 43.7 (39.0-49.1) | 37.4 (33.9-41.2) |
| **Smoking now** |  |  |  |  |  |  |  |  |
| No | 328 | 38.6 (36.5-40.8) | 44.7 (42.0-47.6) | 29.2 (27.7-30.8) | 303 | 52.1 (48.2-56.3) | 75.0 (68.3-82.3) | 49.1 (45.5-53.0) |
| Yes | 56 | 45.1 (40.7-50.1) | 41.9 (37.2-47.2) | 45.6 (41.3-50.3) | 61 | 71.9 (62.7-82.4) | 90.2 (76.5-106.3) | 70.5 (61.6-80.7) |
| **Household air pollution-related factors** |  |  |  |  |  |  |  |  |
| **Household size** |  |  |  |  |  |  |  |  |
| ≤ 4 persons | 196 | 39.6 (37.7-41.7) | 42.6 (40.2-45.2) | 34.2 (32.6-35.9) | 190 | 55.5 (51.8-59.5) | 78.9 (72.6-85.8) | 55.9 (52.2-59.9) |
| > 4 persons | 188 | 41.2 (39.0-43.5) | 45.3 (42.6-48.1) | 32.0 (30.5-33.7) | 174 | 60.0 (55.8-64.5) | 79.9 (73.3-87.2) | 53.8 (50.1-57.8) |
| **Cooking frequency** |  |  |  |  |  |  |  |  |
| Daily | 301 | 41.4 (39.1-43.8) | 43.7 (41.0-46.6) | 34.4 (32.7-36.3) | 280 | 59.1 (54.6-64.0) | 82.4 (74.9-90.6) | 55.3 (51.1-59.8) |
| Daily but not personal/ infrequent | 83 | 38.6 (35.7-41.8) | 44.2 (40.3-48.3) | 31.1 (28.8-33.5) | 84 | 55.2 (49.5-61.5) | 74.9 (65.7-85.3) | 54.4 (48.8-60.6) |
| **Smoky home while cooking** |  |  |  |  |  |  |  |  |
| No | 222 | 37.2 (35.2-39.2) | 40.9 (38.5-43.5) | 32.4 (30.8-34.1) | 196 | 45.7 (42.4-49.1) | 61.8 (56.6-67.6) | 46.8 (43.4-50.3) |
| Yes | 120 | 46.1 (43.0-49.3) | 53.7 (49.7-58.0) | 35.1 (32.9-37.4) | 127 | 74.9 (68.7-81.6) | 119.5 (107.7-132.5) | 67.7 (62.1-73.8) |
| **Smoky home while heating** |  |  |  |  |  |  |  |  |
| No | 163 | 44.3 (41.8-47.0) | 44.1 (41.3-47.2) | 34.0 (32.1-35.9) | 151 | 55.3 (51.2-59.8) | 72.6 (66.2-79.6) | 54.2 (50.2-58.4) |
| Yes | 85 | 39.9 (36.8-43.1) | 51.4 (46.9-56.4) | 29.5 (27.4-31.8) | 101 | 82.0 (74.9-89.7) | 127.9 (114.8-142.4) | 72.5 (66.4-79.2) |

\* Number of subjects. † Only adjusted for sex. ‡ Only adjusted for age.

### 3.2 PM_2.5_ levels by fuel use patterns

Compared across primary cooking and heating fuel combinations, solid fuel users had ∼90% higher personal and kitchen PM_2.5_ levels than those who used clean fuels or did not cook or heat (**Figure 1A**). Personal and household PM_2.5_ levels were significantly (∼75%) higher in winter among solid fuel users, but less so for other participants (∼20%), whilst community levels were 2-3 times higher across different fuel combinations. Broadly similar patterns were observed when examining cooking and heating fuel combinations separately (**Figures 1B and 1C**), but there was a more obvious gradient of exposure at personal and kitchen levels across cooking fuel combinations (as opposed to heating fuel), from the lowest in non-cooking households (winter: ∼50 µg/m_3_) to the highest in solid fuel users (∼115µg/m_3_). A sensitivity analysis restricted to participants who personally cooked regularly showed similar patterns (**eFigure 3**). Consistently, participants who had used solid fuels for cooking or heating had the highest weighted average annual PM_2.5_ exposure at the personal (77.8 [71.1-85.2] µg/m_3_; ∼90% higher), kitchen (103.7 [91.5-117.6] µg/m_3_; ∼130% higher), and living room (62.0 [57.1-67.4] µg/m_3_; ∼65% higher) levels, compared to those who reported using clean fuels or not cooking or heating (**Table 2**). There was no material difference in annual community PM_2.5_ levels by these groups. Similar patterns were observed when examining by cooking and heating fuel combinations separately.

**Figure 1.**
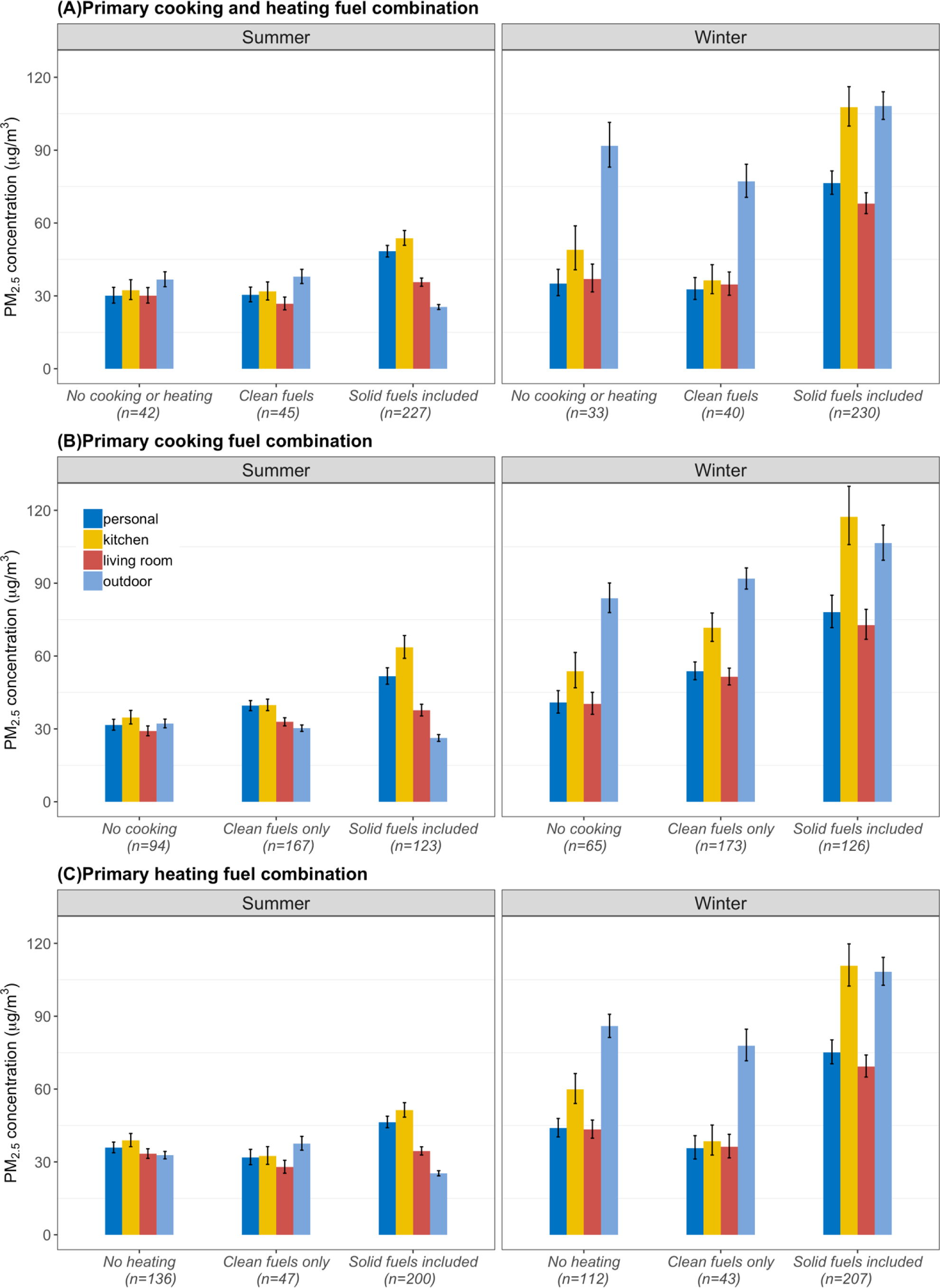
Age- and sex-adjusted geometric mean PM_2.5_ concentrations (μg/m^3^) recorded in the personal, kitchen, living room, and community monitors by season and the combination of primary cooking and heating fuels. Each vertical bar represents adjusted geometric means of each microenvironment by exposure groups, with vertical black lines showing the corresponding 95% confidence intervals (CIs). Non-overlapping CIs between bars indicate statistically significant difference. From left to right the four bars in each group are personal, kitchen, living room, and community PM_2.5_ levels. Participants reporting using unspecified “other” fuels for heating were excluded due to small sample size (N_summer_ = 1; N_winter_ = 2).

**Table 2.** Age- and sex-adjusted estimated annual mean PM_2·5_ exposure levels (μg/m^3^) for the personal, kitchen, living room, and community environments by cooking and heating fuel category *

| **Cooking and heating fuel category** | **Personal** | **Kitchen** | **Living room** | **Community** |
| --- | --- | --- | --- | --- |
| **Primary cooking fuel combination** |  |  |  |  |
| No cooking (n=71) | 45.2 (39.3-52.1) | 60.6 (49.9-73.6) | 42.6 (37.7-48.3) | 55.0 (49.7-60.9) |
| Clean fuels only (n=135) | 57.8 (52.3-63.9) | 69.5 (60.5-79.9) | 49.9 (45.7-54.6) | 56.3 (52.1-60.8) |
| Solid fuels included (n=101) | 86.7 (76.5-98.3) | 121.6 (102.3-144.6) | 68.9 (61.7-76.9) | 63.1 (57.1-69.8) |
| **Primary heating fuel combination** |  |  |  |  |
| No heating (n=98) | 50.2 (44.4-56.8) | 65.9 (55.7-77.9) | 47.2 (42.4-52.7) | 56.3 (51.8-61.2) |
| Clean fuels only (n=39) | 43.4 (36.1-52.2) | 44.5 (34.6-57.2) | 39.3 (33.4-46.3) | 53.7 (47.7-60.6) |
| Solid fuels included (n=169) | 76.3 (69.2-84.2) | 103.5 (90.6-118.3) | 61.3 (56.3-66.9) | 61.4 (56.3-66.9) |
| **Primary cooking and heating fuel combination** |  |  |  |  |
| No cooking or heating (n=29) | 38.4 (31.0-47.6) | 50.1 (37.2-67.6) | 41.1 (33.8-50.1) | 60.0 (51.2-70.3) |
| Clean fuels only (n=37) | 40.9 (34.2-48.9) | 43.5 (33.9-55.8) | 37.9 (32.1-44.7) | 53.2 (46.7-60.7) |
| Solid fuels included (n=189) | 77.8 (71.1-85.2) | 103.7 (91.5-117.6) | 62.0 (57.1-67.4) | 61.7 (56.4-67.6) |

\* Annual mean level was estimated using the regional temperature data from 2005-2017, the number of months with average temperature <10 degrees are 3/12 in Suzhou, 5/12 in Gansu, 4/12 in Henan. Using the same data, the proportion of days with ≤10 degrees daily average temperature are 1233/4717 (26.1%) in Suzhou, 1921/4717 (40.7%) in Gansu, 1524/4717 (32.7%) in Henan.

When examining the aggregated diurnal PM_2.5_ patterns by cooking fuel combinations, we observed major peaks at around noon and evening time for both solid fuel and clean fuel users (less so for non-cooking households), and these peaks were substantially higher in solid fuel users, up to ∼600 and ∼1200µg/m_3_ in summer and winter, respectively (**Figure 2A-C**). A small morning peak (∼08:00) was also found in the kitchen in winter (**Figure 2B**). Interestingly, personal, kitchen, and living room PM_2.5_ levels were considerably higher most of the time among clean fuel users than non-cooking households. Solid fuel users appeared to have the lowest community PM_2.5_ exposure in summer, but highest in winter, with broadly concordant diurnal variations for all three fuel use categories (**Figure 2D**).

**Figure 2.**
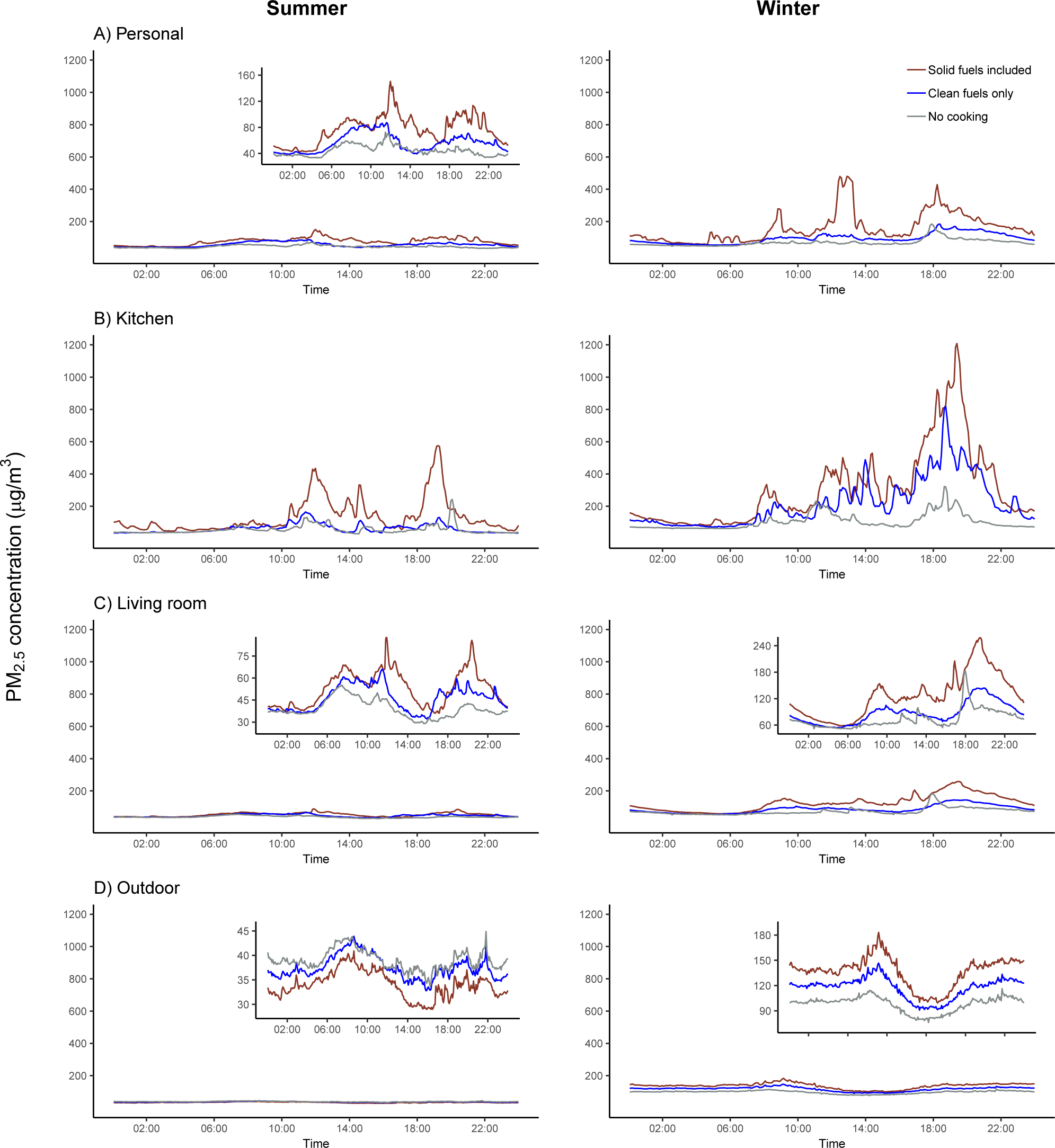
24-hour average time-series plots for PM_2.5_ concentrations (μg/m^3^) recorded in the personal, kitchen, living room, and community monitors by season and primary cooking fuel combinations. There were 123 (13761 person-hour), 167 (18663 person-hour) and 94 (10607 person-hour) subjects for the “Solid fuels included”, “Clean fuels” and “No cooking” group in summer, respectively; There were 126 (13174 person-hour), 173 (17956 person-hour) and 65 (6819 person-hour) subjects for the “Solid fuels included”, “Clean fuels” and “No cooking” group in winter, respectively. Smaller plots nested within panels are “zoom-in” version of the corresponding plot, as the use of a universal y-axis limit up to 1200 with reference to the kitchen exposure levels impaired the readability of those plots.

The differences in diurnal exposure levels by heating fuel combinations were largely similar to those by cooking fuels in summer, but in winter the exposure levels in individuals who did not have heating were just slightly lower than the solid fuel users but much higher than clean fuel users (**Figure 3**).

**Figure 3.**
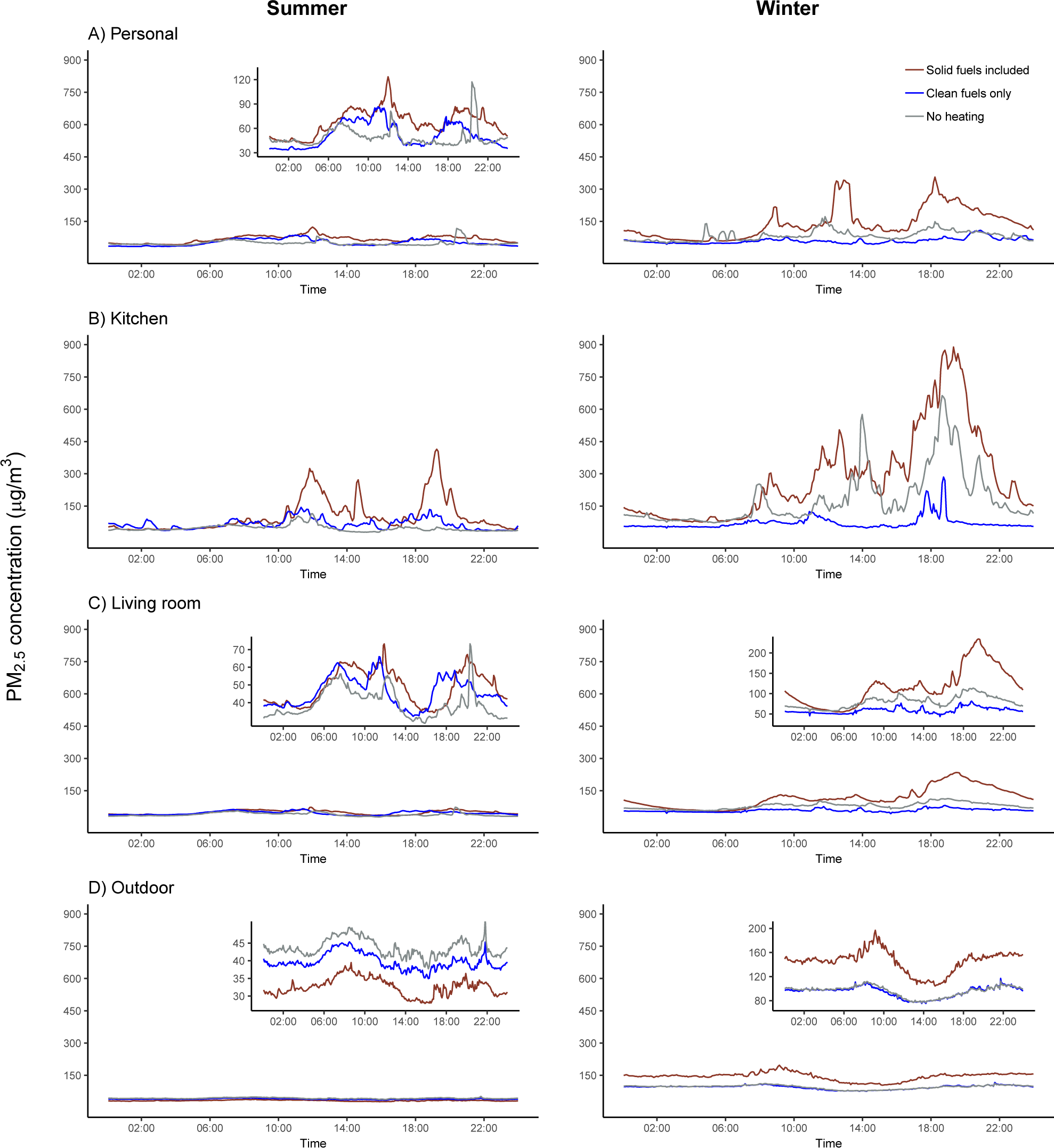
24-hour average time-series plots for PM_2.5_ concentrations (μg/m^3^) recorded in the personal, kitchen, living room, and community monitors by season and primary heating fuel combinations. There were 200 (22539 person-hours), 47 (5226 person-hours) and 136 (15147 person-hours) subjects for the “Solid fuels included”, “Clean only” and “No heating” group in summer, respectively; There were 207 (21772 person-hours), 43 (4261 person-hours) and 112 (11701 person-hours) subjects for the “Solid fuels included”, “Clean only” and “No heating” group in winter, respectively. Smaller plots nested within panels are “zoom-in” version of the corresponding plot, as the use of a universal y-axis limit up to 1200 with reference to the kitchen exposure levels impaired the readability of those plots.

### 3.3 PM_2.5_ exposure models and inter-spatial correlation

We found moderate correlation between measured PM_2.5_ at personal, living room (r: 0.64-0.66), and kitchen (0.52-0.59) levels, whilst the correlation of personal and household levels with community levels was weaker, especially in summer (r_range_summer_: 0.15-0.34; r_range_winter_: 0.41-0.55) (**Figure 4**). Stratified by cooking and heating fuel combinations, we found the highest correlation of personal and household with community levels among those reporting no cooking or heating (r_range_summer_: 0.52-0.65; r_range_winter_: 0.55-0.62), and weakest among those who used solid fuels (r_range_summer_: 0.11-0.31; r_range_winter_: 0.29-0.52) (**eFigures 4A-C**).

**Figure 4.**
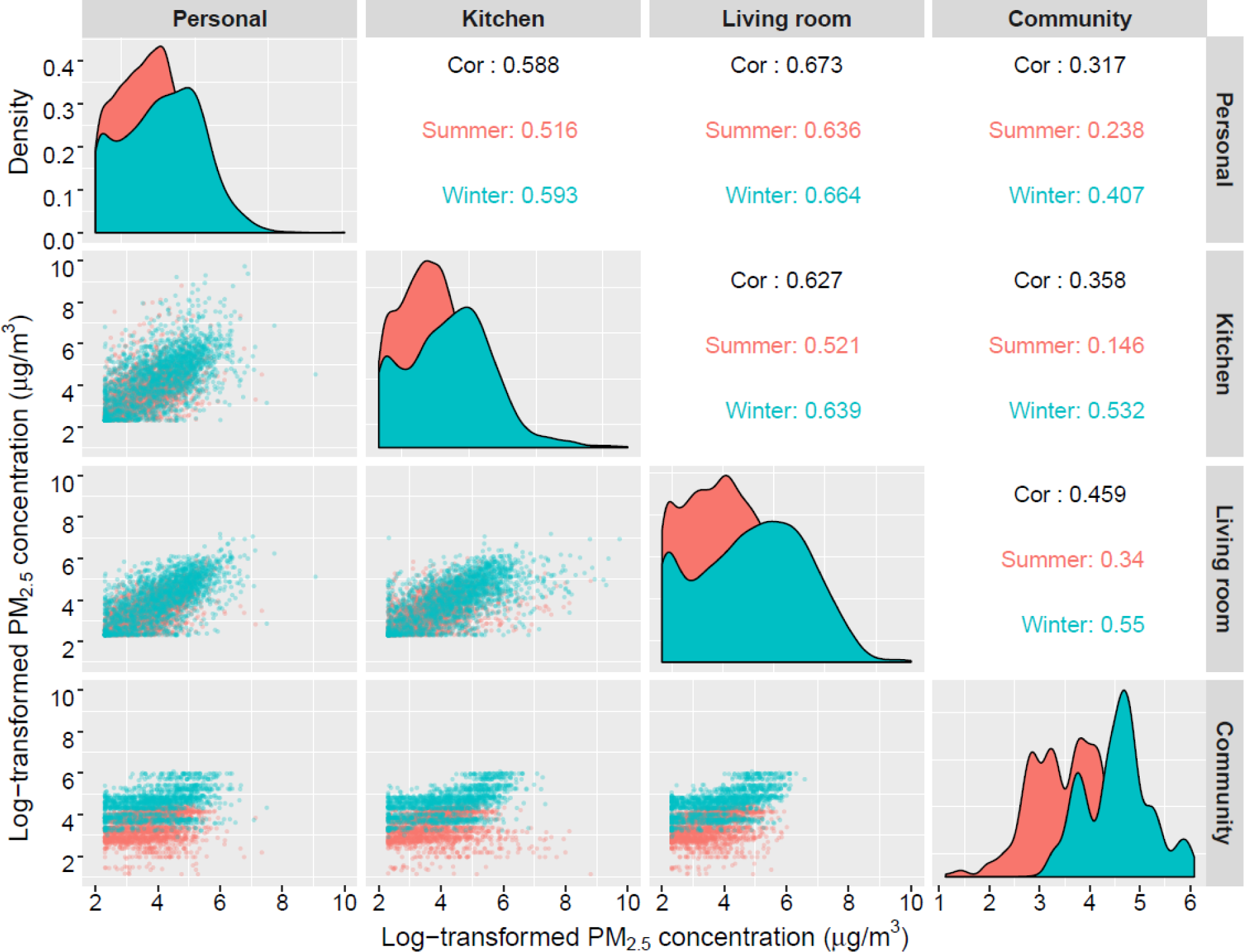
The correlation matrix between the log-transformed concentrations of PM_2.5_ at personal, kitchen, living room and community levels. Red area under curves and dots are summer data; blue area under curves and dots are winter data; black numbers in boxes are overall Spearman correlation coefficient; red and blue numbers are summer- and winter-specific correlation.

## 4. Discussion

We reported integrated and time-resolved PM_2.5_ levels at personal, household (kitchen and living room), and community environments by cooking and heating fuel combinations and other key characteristics in ∼360 adults from one urban and two rural areas of China. Solid fuel use for cooking and heating was associated with significantly higher estimated annual PM_2.5_ exposure at both personal and household levels, with personal PM_2.5_ exposure at ∼3 times and 5 times the World Health Organization 24-hour Air Quality Guidelines (WHO AQG) level (15µg/m_3_) in summer and winter, respectively, and an estimated annual personal exposure at over 15 times of the WHO annual AQG level (5µg/m^3^)^20^. The PM_2.5_ levels across all microenvironments were higher in winter than summer, with ∼2-3 times higher community levels regardless of fuel use. Time-resolved data showed vast inter-and intra-personal variability in PM_2.5_ exposure within and across seasons, with remarkably high exposure (5-min moving-average up to 1200µg/m_3_) recorded in typical cooking times (∼2-4 hours per day), most notably in the kitchen but also personal monitors among solid fuel users.

Previous studies assessing exposures to air pollution were highly heterogeneous in settings, sample size, prevalent fuel types, and recorded PM_2.5_ levels,^6–14^ but there has been broadly consistent evidence that solid fuel use for cooking was associated with higher personal and kitchen PM_2.5_ levels as reported in our study. For logistical and technical reasons, most previous studies primarily measured kitchen PM_2.5_,^6–14, 21^ while some had parallel measurements of personal_6,7,14_ or ambient_10,14,21_ exposure, with most personal measurements done in a subset of participants. Notably, the largest single sample (n=998; ∼48,000 person-hours) of personal PM_2.5_ measurements (alongside kitchen measurements in 2,541 households) came from the PURE-Air study focussing on cooking fuel in rural areas across eight countries.^6^ With 48-hour integrated PM_2.5_ measurements, they found lower PM_2.5_ levels by cooking fuel types moving up the traditional ‘energy ladder’ (i.e. from heavily polluting biomass to coal, then to gas and electricity),_22_ but they also found substantial heterogeneity within each solid fuel category and between countries (e.g. kitchen PM_2.5_ for primary wood use was 50 [45-55] µg/m^3^ in China and 105 [96-116] µg/m_3_ in India), possibly due to varying fuel use behaviour or infrastructure, chemical constituents of fuels, and different climate conditions. With the parallel and repeated time-resolved assessment of personal, kitchen, living room (∼81,000 person-hours for each measure), and community (∼67,000 person-hours) level PM_2.5_ in summer and winter, we provided further insight into the complex relationships between fuel use behaviour and PM_2.5_ levels across the personal-household-community exposure spectrum.

Consistent with the few existing studies that ascertained multiple fuel use,_6,10,14_ we showed that fuel stacking was common in rural China, and mixed use of solid and clean fuels was associated with substantially elevated PM_2.5_ exposure, especially in winter. As fuel stacking is increasingly common in many developing economies, this highlights the importance of capturing usage information beyond a single, primary fuel type, in order to more accurately assess household air pollution exposure and the associated disease burden. On the other hand, until recently, both researchers and policymakers have largely overlooked heating as a major contributor of air pollution._18,23-25_ Our findings add to the previous field measurement studies,_26-28_ showing solid fuels for heating to be associated with 76-125% higher PM_2.5_ exposure at personal and household levels in winter. It may seem counterintuitive to observe a higher level in the kitchen than in the living room, but previous studies have noted poorer ventilation in winter and that solid fuel users may stay in the kitchen longer to get warmth from the cookstove to save fuel._22_ In line with a slower rate of modernisation of heating (versus cooking) fuel in China (as in many other LMICs),_18,24_ about 50% of CKB-Air participants who had used clean fuels for cooking still relied on solid fuels for heating. Adding to the complexity, the lack of heating in rural China was associated with a lower socioeconomic status and greater likelihood of using solid fuels for cooking compared to clean fuel users._18_ This, together with the likely reduced ventilation (to keep warm) in winter time, may explain the considerably (∼30%) higher personal and household PM_2.5_ levels (in both seasons) among our participants who reported ‘no heating’, compared with the clean heating fuel users.

While the community PM_2.5_ level was markedly higher in winter regardless of fuel use categories, there was an interesting contrast that solid fuel users had the lowest community PM_2.5_ in summer, but the highest in winter. The ‘winter smog’ phenomenon in densely populated (often urban) areas of China is well-documented, as increased energy consumption, reliance on coal-fired power plants, and meteorological factors (e.g. temperature inversion) drive heightened regional ambient air pollution._29_ On the other hand, most solid fuel users resided in rural areas with lower population and vehicle density, which tend to be associated with lower ambient air pollution. In winter, however, the intensive use of solid fuels for heating (most participants reported heating throughout the day) could result in major rise of neighbourhood PM_2.5_ in addition to regional ambient air pollution, as supported by previous studies._14,30_ It is also worth noting that the increase in personal and household levels was much higher in solid fuel users than in other participants, whose personal and household levels were less than 50% of the community levels.

As in many previous studies_14,21_ we observed relatively weak correlation between personal and community PM_2.5_ levels, which poses challenges to the previous disease burden estimates for ambient air pollution in LMICs based mainly on epidemiological studies using modelled ambient levels without accounting for inter-spatial variability and people’s time spent indoors (typically 70-80%)._31_ This may be less problematic in HICs with relatively low exposure from non-ambient sources, although the re-emergence of wood-fire heating may raise concern._32_ The relatively strong correlation (0.52-0.66) between personal and household measurements is consistent with previous evidence (e.g. PURE-Air: person-to-kitchen correlation = 0.69). Our evidence adds further support for more granular household measurements along with housing characteristics questionnaires, simple personal GPS trackers, and advanced ambient air pollution modelling approaches_33,34_ to better approximate personal exposure in large-scale epidemiological studies. More in-depth modelling analysis on our data will generate further insight for better exposure approximation in future studies.

Our time-resolved data also illustrated the remarkable short-term intra- and inter-personal variability in PM_2.5_ exposure even within each fuel use category. The diurnal patterns of kitchen PM_2.5_ appeared consistent with the previously reported time-activity patterns in CKB-Air,_15_ such as the exposure peaks (averaged twice in summer; 3 times in winter) at typical meal times. Furthermore, we observed stronger and longer-lasting evening peaks of personal and household levels among individuals who used solid fuels in winter, which is consistent with typical space heating practices with reduced ventilation at night. The vast diurnal variations, with personal PM_2.5_ exposure as high as 400µg/m^3^ and as low as 10µg/m_3_, lead to the question of whether and how long-term average exposure could compare to an accumulation of repeated bursts of extreme exposure in relation to disease development risk._35_ The mystery might be solved by the increasing availability of more refined air pollution data, the use of chamber studies, and the emerging multi-omics technologies that facilitate a better understanding of the toxicology and pathophysiology.

CKB-Air offers one of the most detailed parallel and repeated seasonal assessments of personal, household, and community level PM_2.5_ with one of the largest time-resolved datasets (up to ∼80,000 person-hours per microenvironment). Moreover, we assessed not only the role of parallel fuel use for cooking but also for heating on both average and time-resolved PM_2.5_ exposure, shedding light on the complexity of fuel use behaviour and PM_2.5_ exposure. However, several limitations warrant discussion. First, despite the relatively large amount of data captured, the number of participants representing each fuel use combination beyond the aggregated categories on solid versus clean fuels was small.

Also, the large inter-and intra-personal variability means that we could not reliably estimate PM_2.5_ levels by >10 different fuel combinations captured. Second, unlike some previous studies that used gold-standard gravimetric samplers in measuring integrated PM_2.5_ exposure,_6_ we used a nephelometer in order to obtain detailed time-resolved data. Despite the field- and lab-based validation and calibration, our instruments inevitably entailed measurement error, but this should not result in major biases that would affect the applicability in epidemiological studies. Third, we assessed community PM_2.5_ at a single location, and we lacked pairwise data of street and regional levels. Fourth, the study sample was recruited via convenient sampling from three purposively selected areas in China, so the estimated exposure levels would not be generalisable to China or other populations.

## 5. Conclusions

This study has demonstrated the feasibility and value of collecting detailed air pollution exposure measurement data to capture intra- and inter-personal variations over short (weekly) and medium (seasonal) term, in rural and urban China. Most notably, the individuals who used solid fuels for cooking or heating were found to have annual personal PM_2.5_ exposure over 15 times higher than the latest WHO AQG. The relatively weak correlation of personal with community PM_2.5_, in contrast to the stronger correlation between personal and household levels, supports the use of reliable, low-cost household static monitors in improving personal air pollution exposure assessment in large-scale epidemiological studies. Our findings underscores the complexity of air pollution exposure and the need for cross-disciplinary investigation involving exposure science, toxicology, epidemiology and statistics.

## Data Availability

The China Kadoorie Biobank (CKB) is a global resource for the investigation of lifestyle, environmental, blood biochemical and genetic factors as determinants of common diseases. The CKB study group is committed to making the cohort data available to the scientific community in China, the UK and worldwide to advance knowledge about the causes, prevention and treatment of disease. For detailed information on what data is currently available to open access users and how to apply for it, visit: http://www.ckbiobank.org/site/Data+Access.
Researchers who are interested in obtaining the raw data from the China Kadoorie Biobank study that underlines this paper should contact. A research proposal will be requested to ensure that any analysis is performed by bona fide researchers.

## Acknowledgements

The chief acknowledgment is to the participants, the project staff, and the China National Centre for Disease Control and Prevention (CDC) and its regional offices for assisting with the fieldwork. We thank Judith Mackay in Hong Kong; Yu Wang, Gonghuan Yang, Zhengfu Qiang, Lin Feng, Maigeng Zhou, Wenhua Zhao, and Yan Zhang in China CDC; Lingzhi Kong, Xiucheng Yu, and Kun Li in the Chinese Ministry of Health; and Garry Lancaster, Sarah Clark, Martin Radley, Mike Hill, Hongchao Pan, and Jill Boreham in the CTSU, Oxford, for assisting with the design, planning, organization, and conduct of the study.

The CKB baseline survey and the first re-survey were supported by the Kadoorie Charitable Foundation in Hong Kong. The long-term follow-up has been supported by Wellcome grants to Oxford University (212946/Z/18/Z, 202922/Z/16/Z, 104085/Z/14/Z, 088158/Z/09/Z) and grants from the National Natural Science Foundation of China (82192900, 82192901, 82192904) and from the National Key Research and Development Program of China (2016YFC0900500).The UK Medical Research Council (MC_UU_00017/1,MC_UU_12026/2, MC_U137686851), Cancer Research UK (C16077/A29186; C500/A16896) and the British Heart Foundation (CH/1996001/9454), provide core funding to the Clinical Trial Service Unit and Epidemiological Studies Unit at Oxford University for the project. KHC acknowledges support from the BHF Centre of Research Excellence, University of Oxford (RE/18/3/34214). AD is supported by the Wellcome Trust (223100/Z21/Z). The CKB-Air study was supported by a UK Medical Research Council: Global Challenges Research Fund – Foundation Award (Ref MR/P025080/1) and a Nuffield Department of Population Health Pump-priming Award.

## Open Access Statement

This research was funded in whole, or in part, by the Wellcome Trust [Grant number 212946/Z/18/Z, 202922/Z/16/Z, 104085/Z/14/Z, 088158/Z/09/Z, 223100/Z21/Z]. For the purpose of Open Access, the author has applied a CC-BY public copyright licence to any Author Accepted Manuscript version arising from this submission.

## Data Access Statement

The China Kadoorie Biobank (CKB) is a global resource for the investigation of lifestyle, environmental, blood biochemical and genetic factors as determinants of common diseases. The CKB study group is committed to making the cohort data available to the scientific community in China, the UK and worldwide to advance knowledge about the causes, prevention and treatment of disease. For detailed information on what data is currently available to open access users and how to apply for it, visit: http://www.ckbiobank.org/site/Data+Access.

Researchers who are interested in obtaining the raw data from the China Kadoorie Biobank study that underlines this paper should contact. A research proposal will be requested to ensure that any analysis is performed by bona fide researchers.

## Declaration of interest

The authors declare no known conflict of interest.

## CRediT authorship contribution statement

Ka Hung Chan: Conceptualization, Methodology, Software, Formal analysis, Investigation, Resources, Data curation, Writing – original draft, Writing – review & editing, Visualization, Project administration, Supervision. Xi Xia: Methodology, Software, Formal analysis, Investigation, Writing – review & editing, Visualization. Cong Liu, Haidong Kan: Methodology, Resources, Data curation, Writing – review & editing. Aiden Doherty: Methodology, Investigation, Writing – review & editing. Hung Lam Steve Yim: Methodology, Resources, Writing – review & editing. Neil Wright, Christiana Kartsonaki: Methodology, Software, Data curation, Writing – review & editing. Xiaoming Yang, Rebecca Stevens: Resources, Data curation, Software. Xiaoyu Chang: Resources, Project administration. Canqing Yu, Jun Lv, Liming Li: Resources, Project administration, Funding acquisition. Kin-Fai Ho: Conceptualization, Methodology, Resources, Writing – review & editing, Supervision. Kin Bong Hubert Lam, Zhengming Chen: Conceptualization, Methodology, Resources, Writing – review & editing, Project administration, Supervision, Funding acquisition.

## Notes

### Competing Interest Statement

The authors have declared no competing interest.

