## Supplementary materials for "Characterising personal, household, and community PM2.5 exposure in one urban and two rural communities in China"

**Text S1. Members of the China Kadoorie Biobank collaborative group**

**International Steering Committee:** Junshi Chen, Zhengming Chen (PI), Robert Clarke, Rory Collins, Liming Li (PI), Chen Wang, Jun Lv, Richard Peto, Robin Walters.

**International Co-ordinating Centre, Oxford:** Daniel Avery, Maxim Barnard, Derrick Bennett, Ruth Boxall, Sushila Burgess, Ka Hung Chan, Yiping Chen, Zhengming Chen, Johnathan Clarke; Robert Clarke, Huaidong Du, Ahmed Edris Mohamed, Hannah Fry, Simon Gilbert, Pek Kei Im, Andri Iona, Maria Kakkoura, Christiana Kartsonaki, Hubert Lam, Kuang Lin, James Liu, Mohsen Mazidi, Iona Millwood, Sam Morris, Qunhua Nie, Alfred Pozarickij, Paul Ryder, Saredo Said, Dan Schmidt, Becky Stevens, Iain Turnbull, Robin Walters, Baihan Wang, Lin Wang, Neil Wright, Ling Yang, Xiaoming Yang, Pang Yao.

**National Co-ordinating Centre, Beijing:** Xiao Han, Can Hou, Qingmei Xia, Chao Liu, Jun Lv, Pei Pei, Dianjianyi Sun, Canqing Yu.

**10 Regional Co-ordinating Centres:**

**Guangxi** Provincial CDC: Naying Chen, Duo Liu, Zhenzhu Tang. Liuzhou CDC: Ningyu Chen, Qilian Jiang, Jian Lan, Mingqiang Li, Yun Liu, Fanwen Meng, Jinhuai Meng, Rong Pan, Yulu Qin, Ping Wang, Sisi Wang, Liuping Wei, Liyuan Zhou. **Gansu** Provincial CDC: Caixia Dong, Pengfei Ge, Xiaolan Ren. Maiji CDC: Zhongxiao Li, Enke Mao, Tao Wang, Hui Zhang, Xi Zhang. **Hainan** Provincial CDC: Jinyan Chen, Ximin Hu, Xiaohuan Wang. Meilan CDC: Zhendong Guo, Huimei Li, Yilei Li, Min Weng, Shukuan Wu. **Heilongjiang** Provincial CDC: Shichun Yan, Mingyuan Zou, Xue Zhou. Nangang CDC: Ziyan Guo, Quan Kang, Yanjie Li, Bo Yu, Qinai Xu. **Henan** Provincial CDC: Liang Chang, Lei Fan, Shixian Feng, Ding Zhang, Gang Zhou. Huixian CDC: Yulian Gao, Tianyou He, Pan He, Chen Hu, Huarong Sun, Xukui Zhang. **Hunan** Provincial CDC: Biyun Chen, Zhongxi Fu, Yuelong Huang, Huilin Liu, Qiaohua Xu, Li Yin. Liuyang CDC: Huajun Long, Xin Xu, Hao Zhang, Libo Zhang. **Jiangsu** Provincial CDC: Jian Su, Ran Tao, Ming Wu, Jie Yang, Jinyi Zhou, Yonglin Zhou. Suzhou CDC: Yihe Hu, Yujie Hua, Jianrong Jin, Fang Liu, Jingchao Liu, Yan Lu, Liangcai Ma, Aiyu Tang, Jun Zhang. **Qingdao** CDC: Liang Cheng, Ranran Du, Ruqin Gao, Feifei Li, Shanpeng Li, Yongmei Liu, Feng Ning, Zengchang Pang, Xiaohui Sun, Xiaocao Tian, Shaojie Wang, Yaoming Zhai, Hua Zhang, Licang CDC: Wei Hou, Silu Lv, Junzheng Wang. **Sichuan** Provincial CDC: Xiaofang Chen, Xianping Wu, Ningmei Zhang, Xiaoyu Chang. Pengzhou CDC: Xiaofang Chen, Jianguo Li, Jiaqiu Liu, Guojin Luo, Qiang Sun, Xunfu Zhong. **Zhejiang** Provincial CDC: Weiwei Gong, Ruying Hu, Hao Wang,Meng Wang, Min Yu. Tongxiang CDC: Lingli Chen, Qijun Gu, Dongxia Pan， Chunmei Wang, Kaixu Xie, Xiaoyi Zhang.

**Text S2. Household questionnaire of CKB-Air**

### Household survey questionnaire (for cool season only)

**Note: All questions after Q1.5 in this document are either new or different from the questions included in the previous CKB surveys, unless otherwise specified.**

#### Section 1: Background information (This section is for ParticipantsDetails : Grey is not collected in SINPUT)

**1.3** Name: ________________, **Sex:** Male ☐ Female ☐

**1.4 Date of birth:** ☐☐☐☐ Year ☐☐ Month ☐☐ Day

**1.5** National ID: ☐☐☐☐☐☐☐☐☐☐☐☐☐☐☐☐☐☐

---

_

**1.6 What is your current occupation?**

- ☐ Agriculture & related workers
- ☐ Factory worker
- ☐ Administrator / manager
- ☐ Professional / technical
- ☐ Sales & service workers
- ☐ Retired
- ☐ House wife / husband
- ☐ Self-employed
- ☐ Unemployed
- ☐ Other or not stated

**1.7 How many people live together as a family in the household?** _____persons (same as RS2 Q1.8)

**1.8 Type of dwelling:** ☐ Apartment ☐ House

**1.9 What is the total income last year in your household?** (same as RS2 Q1.10)

- ☐ <2,500 yuan
- ☐ 2,500-4,999 yuan
- ☐ 5,000-9,999 yuan
- ☐ 10,000-19,999 yuan
- ☐ 20,000-34,999 yuan
- ☐ 35,000-49,999 yuan
- ☐ 50,000-74,999 yuan
- ☐ 75,000-99,999 yuan
- ☐ ≥100,000 yuan

#### Section 2: Active and passive smoking

**2.1 How often do you smoke tobacco now?** (Comparable to RS2 4.2)

- ☐ Do not smoke now
- ☐ Only occasionally
- ☐ Yes, on most days
- ☐ Yes, daily or almost every day

**2.2 During the past 12 months, how frequently have you been exposed to tobacco smoke from a family member at home?** (i.e. a minimum of 5 consecutive minutes each time) (Modified from 6.2 & 6.2.1)

- ☐ Never } if ticked, go to Q2.3
- ☐ <1 day/week } if ticked, go to Q2.3
- ☐ 1-2 day/week
- ☐ 3-5 day/week
- ☐ 6-7 day/week

**2.2.1 What is the usual duration of your exposure per week?** ☐☐ Hours

**2.3 During the past 12 months, how frequently have you been exposed to other people's tobacco smoke in workplace or public place?** (i.e. a minimum of 5 consecutive minutes each time) (Modified from 6.3 & 6.3.1)

- ☐ Never } if ticked, go to Q3.1
- ☐ <1 day/week } if ticked, go to Q3.1
- ☐ 1-2 day/week
- ☐ 3-5 day/week
- ☐ 6-7 day/week

**2.3.1 What is the usual duration of your exposure per week?** ☐☐ Hours

### Section 3: Cooking related exposure

**3.1 During the past 12 months, how often was cooking done (by anyone) at your home?**

- ☐ Daily or almost every day
- ☐ A few times a week
- ☐ A few times a month
- ☐ Never or rarely → if ticked, go to Section 4 Q4.1

**3.2 During the past 12 months, how often did you cook at home?**

- ☐ Daily or almost every day } if ticked, go to Q3.3.1
- ☐ A few times a week } if ticked, go to Q3.3.1
- ☐ A few times a month
- ☐ Never or rarely

**3.3 Have you ever cooked regularly at home (at least few times a week)?**

☐ Yes ☐ No → if ticked, go to Q3.5

**3.3.1 At about what age did you start cooking regularly at home**? ☐☐ Years

→ If ticked 1st or 2nd box in Q3.2, go to Q3.4

**3.3.2 At about what age did you stop cooking regularly at home?**

☐Since _______(year old) → if answered "Daily or almost every day" or "A few times a week" for **Q3.2**, this question should be skipped.

**3.4 During the time period that you were cooking regularly at home, how much time do you spend in front of the fire/ cookstove/ in the kitchen while cooking per day?** _______ Hours

**3.5 In your household, which cooking fuels are used now, how frequently and how long have they been used (during your lifetime)?** *(Excluding boiling water for drinking or the use of electric rice cooker; tick multiple type of fuel if applicable)*

| | **Used** | | **Frequency of use** | | **Duration used (years)** |
|---|---|---|---|---|---|
| | **Yes** | **No** | **Most meals** | **Sometimes** | *(If < 1 year, enter 0)* |
| Electricity (hob/oven) | ☐ | ☐ | ☐ | ☐ | ________ |
| Natural gas/ town gas/ LPG | ☐ | ☐ | ☐ | ☐ | ________ |
| Biogas | ☐ | ☐ | ☐ | ☐ | ________ |
| Smokeless coal | ☐ | ☐ | ☐ | ☐ | ________ |
| Smoky coal | ☐ | ☐ | ☐ | ☐ | ________ |
| Coalite/ coal brick | ☐ | ☐ | ☐ | ☐ | ________ |
| Charcoal | ☐ | ☐ | ☐ | ☐ | ________ |
| Wood | ☐ | ☐ | ☐ | ☐ | ________ |
| Crop residue | ☐ | ☐ | ☐ | ☐ | ________ |
| Kerosene | ☐ | ☐ | ☐ | ☐ | ________ |
| Solar | ☐ | ☐ | ☐ | ☐ | ________ |
| Other | ☐ | ☐ | ☐ | ☐ | ________ |

_______________________________________________________________

**3.6 Have the cooking fuel in your home now (cool season) changed compared to that was used during the warm period (warm season)?**

☐ Yes ☐ No → if ticked, go to Q3.8

**3.7 Which cooking fuels were used in your home back then and for how long?** *(Excluding boiling water for drinking or the use of electric rice cooker; tick multiple type of fuel if applicable)*

| | **Used** | | **Frequency of use** | | **Duration used (years)** |
|---|---|---|---|---|---|
| | **Yes** | **No** | **Most meals** | **Sometimes** | *(If < 1 year, enter 0)* |
| Electricity (hob/oven) | ☐ | ☐ | ☐ | ☐ | ________ |
| Natural gas/ town gas/ LPG | ☐ | ☐ | ☐ | ☐ | ________ |
| Biogas | ☐ | ☐ | ☐ | ☐ | ________ |
| Smokeless coal | ☐ | ☐ | ☐ | ☐ | ________ |
| Smoky coal | ☐ | ☐ | ☐ | ☐ | ________ |
| Coalite/ coal brick | ☐ | ☐ | ☐ | ☐ | ________ |
| Charcoal | ☐ | ☐ | ☐ | ☐ | ________ |
| Wood | ☐ | ☐ | ☐ | ☐ | ________ |
| Crop residue | ☐ | ☐ | ☐ | ☐ | ________ |

| | | | | | |
|---|---|---|---|---|---|
| Kerosene | ☐ | ☐ | ☐ | ☐ | ______ |
| Solar | ☐ | ☐ | ☐ | ☐ | ______ |
| Other | ☐ | ☐ | ☐ | ☐ | ______ |

_

**3.8 Where is cooking in your home usually done?**

☐ Indoors ☐ Outdoors → if ticked, go to Q4.1

**3.9 How often is the kitchen window(s) opened when cooking is done in your home?**

☐ Always ☐ Sometimes ☐ Rarely/ never/no window in the kitchen

**3.10 Does your kitchen have a chimney / extractor fan/hood?**

☐ Yes ☐ No

**3.11 Does the inside of your kitchen tend to be smoky when cooking?**

☐ Always ☐ Sometimes ☐ Rarely/ never

### Section 4: Heating related exposure

**4.1 In winter, how frequently do you normally heat your home?**

☐ Daily or almost every day

☐ A few times a week

☐ A few times a month

☐ Never/No heating →if ticked, go to Q4.7

**4.2 How long do you usually use heating on a typical day in winter?** (*consider < 1 hour as 1 hour*) ______Hours

**4.3 Is your residence heated mainly by central heating in winter?** ☐ Yes ☐ No

**4.4 What heating fuels do you use in winter and for how long on a typical day, and how long have you been using them (during your lifetime)?** *(tick multiple type of fuel if applicable)*

| | **Used** | | **Duration of use (hour) in a typical day** *(to the nearest 0.5 hr)* | **Duration used (years)** *(If < 1 year, enter 0)* |
|---|---|---|---|---|
| | **Yes** | **No** | | |
| Electricity | ☐ | ☐ | ______ | ______ |
| Natural gas/ town gas/ LPG | ☐ | ☐ | ______ | ______ |
| Biogas | ☐ | ☐ | ______ | ______ |
| Smokeless coal | ☐ | ☐ | ______ | ______ |
| Smoky coal | ☐ | ☐ | ______ | ______ |
| Coalite/ coal brick | ☐ | ☐ | ______ | ______ |

| | | | | |
|---|---|---|---|---|
| Charcoal | ☐ | ☐ | ______ | ______ |
| Wood | ☐ | ☐ | ______ | ______ |
| Crop residue | ☐ | ☐ | ______ | ______ |
| Kerosene | ☐ | ☐ | ______ | ______ |
| Solar | ☐ | ☐ | ______ | ______ |
| Other | ☐ | ☐ | ______ | ______ |

**4.5 Do your (non-electric) heat stoves have a chimney / extractor?**

☐ Yes, all ☐ Yes, but not all ☐ None

**4.6 Does the inside of your home tend to be smoky when you use heating?**

☐ Always ☐ Sometimes ☐ Rarely/ never

**4.7 What is the primary reason of not using heating?** (only for those who answered “No heating” to Q4.1)

☐ No such need ☐ Cannot afford ☐ Inconvenience

### Section 5: Other sources of air pollution

**5.1 Do you keep a stove under slow burning indoors throughout the day (not for heating)?**

☐ Yes, always ☐ Yes, sometimes ☐ No

**5.2 How frequently did you use mosquito coils in summer?**

☐ Daily or almost every day

☐ A few times a week

☐ A few times a month

☐ Never or rarely

**5.3 During the past 12 months, how frequently did you burn incense indoors?**

☐ Daily or almost every day

☐ A few times a week

☐ A few times a month

☐ Never or rarely

**eTable 1. Distribution of flagged $PM_{2.5}$ data in number of rows of 5-mins moving average data points, according to PATS device location and study season**

| Season | Personal | % | Kitchen | % | Living room | % | Flag nature* |
|---|---|---|---|---|---|---|---|
| Both | 1,011,856 | 95.5 | 1,016,624 | 96.0 | 1,036,312 | 97.8 | normal |
| | 34,979 | 3.3 | 20,706 | 2.0 | 18,465 | 1.7 | low |
| | 12,455 | 1.2 | 21,960 | 2.1 | 4,513 | 0.4 | high |
| Summer | 549,135 | 97.1 | 557,038 | 98.5 | 560,630 | 99.1 | normal |
| | 10,760 | 1.9 | 4,775 | 0.8 | 3,906 | 0.7 | low |
| | 5,712 | 1.0 | 3,794 | 0.7 | 1,071 | 0.2 | high |
| Winter | 462,721 | 93.7 | 459,586 | 93.1 | 475,682 | 96.4 | normal |
| | 24,219 | 4.9 | 15,931 | 3.2 | 14,559 | 2.9 | low |
| | 6,743 | 1.4 | 18,166 | 3.7 | 3,442 | 0.7 | high |

*Flag nature regarded as “normal” for data points with no sign of potential error; “low” as having persistently low $PM_{2.5}$ levels; “high” as having persistently high $PM_{2.5}$ levels.

**eTable 2. Total and per participant person-hour of $PM_{2.5}$ data included in the primary analyses, by device location**

| Device Location | Overall (n=748) | | Summer (n=384) | | Winter (n=364) | |
|---|---|---|---|---|---|---|
| | P-hr | Median (Q1, Q3)* | P-hr | Median (Q1, Q3) | P-hr | Median (Q1, Q3) |
| Personal | 80,980 | 208 (119, 231) | 43,031 | 117 (105, 119) | 37,949 | 113 (94, 117) |
| Kitchen | 80,980 | 208 (119, 231) | 43,031 | 117 (105, 119) | 37,949 | 113 (94, 117) |
| Living room | 80,980 | 208 (119, 231) | 43,031 | 117 (105, 119) | 37,949 | 113 (94, 117) |
| Community | 67,326 | 178 (117, 220) | 39,837 | 117 (104, 118) | 27,489 | 112 (93, 117) |

* P-hr: person-hour; Q1: first quartile; Q3: third quartile; note 307 participants had data in both summer and winter, 77 had only summer data, and 57 had only winter data.

**eTable 3: Baseline characteristics of study participants by cooking and heating fuel combinations**

| Characteristics | Cooking fuel combination | | | Heating fuel combination | | |
|---|---|---|---|---|---|---|
| | No cooking | Clean only | Solid included | No heating | Clean only | Solid included |
| **Age-years, mean (SD)** | 57.95 (6.62) | 57.78 (7.02) | 58.97 (5.90) | 59.65 (6.86) | 56.51 (5.65) | 57.62 (6.46) |
| **Female, n (%)** | 73 (77.66) | 110 (65.87) | 104 (84.55) | 101 (74.26) | 32 (68.09) | 153 (76.5) |
| **Region, n (%)** | | | | | | |
| Suzhou (urban) | 60 (63.83) | 74 (44.31) | 1 (0.81) | 95 (69.85) | 40 (85.11) | / |
| Gansu (rural) | 19 (20.21) | 38 (22.75) | 59 (47.97) | 12 (8.82) | 1 (2.13) | 103 (51.5) |
| Henan (rural) | 15 (15.96) | 55 (32.93) | 63 (51.22) | 29 (21.32) | 6 (12.77) | 97 (48.5) |
| **Education, n (%)** | | | | | | |
| No formal education | 15 (15.96) | 33 (19.76) | 49 (39.84) | 38 (27.94) | 3 (6.38) | 56 (28) |
| Primary & middle school | 34 (36.17) | 58 (34.73) | 41 (33.33) | 50 (36.76) | 14 (29.79) | 69 (34.5) |
| Highschool or above | 45 (47.87) | 76 (45.51) | 33 (26.83) | 48 (35.29) | 30 (63.83) | 75 (37.5) |
| **Occupation, n (%)** | | | | | | |
| Agricultural worker | 25 (26.60) | 50 (29.94) | 65 (52.85) | 27 (19.85) | 3 (6.38) | 110 (55) |
| Factory worker | 3 (3.19) | 15 (8.98) | 2 (1.63) | 11 (8.09) | 3 (6.38) | 6 (3) |
| Home-maker | 9 (9.57) | 45 (26.95) | 52 (42.28) | 21 (15.44) | 9 (19.15) | 75 (37.5) |
| Non-manual labour | 1 (1.06) | 6 (3.59) | 2 (1.63) | 4 (2.94) | 1 (2.13) | 4 (2) |
| Self/ un-employed or other | 56 (59.57) | 51 (30.54) | 2 (1.63) | 73 (53.68) | 31 (65.96) | 5 (2.5) |
| **Smoking, n (%)** | 13 (13.83) | 34 (20.36) | 9 (7.32) | 18 (13.24) | 8 (17.02) | 30 (15) |
| **Cooking fuel combination, n (%)** | | | | | | |
| No cooking | / | / | / | 42 (30.88) | 25 (53.19) | 27 (13.5) |
| Clean only | / | / | / | 69 (50.74) | 20 (42.55) | 77 (38.5) |
| Solid included | / | / | / | 25 (18.38) | 2 (4.26) | 96 (48) |
| **Smoky home while cooking, n (%)** | / | 33 (25.98) | 66 (54.10) | 21 (17.50) | 3 (6.82) | 96 (54.24) |
| **Heating fuel combination, n (%)** | | | | | | |
| No heating | 42 (44.68) | 69 (41.32) | 25 (20.33) | / | / | / |
| Clean only | 25 (26.60) | 20 (11.98) | 2 (1.63) | / | / | / |
| Solid included | 27 (28.72) | 1 (0.60) | 96 (78.05) | / | / | / |
| **Smoky home while heating, n (%)** | 17 (32.69) | 24 (24.49) | 44 (44.90) | / | 1 (2.13) | 84 (42.00) |

**eFigure 1. Study areas of the China Kadoorie Biobank Cohort Study***

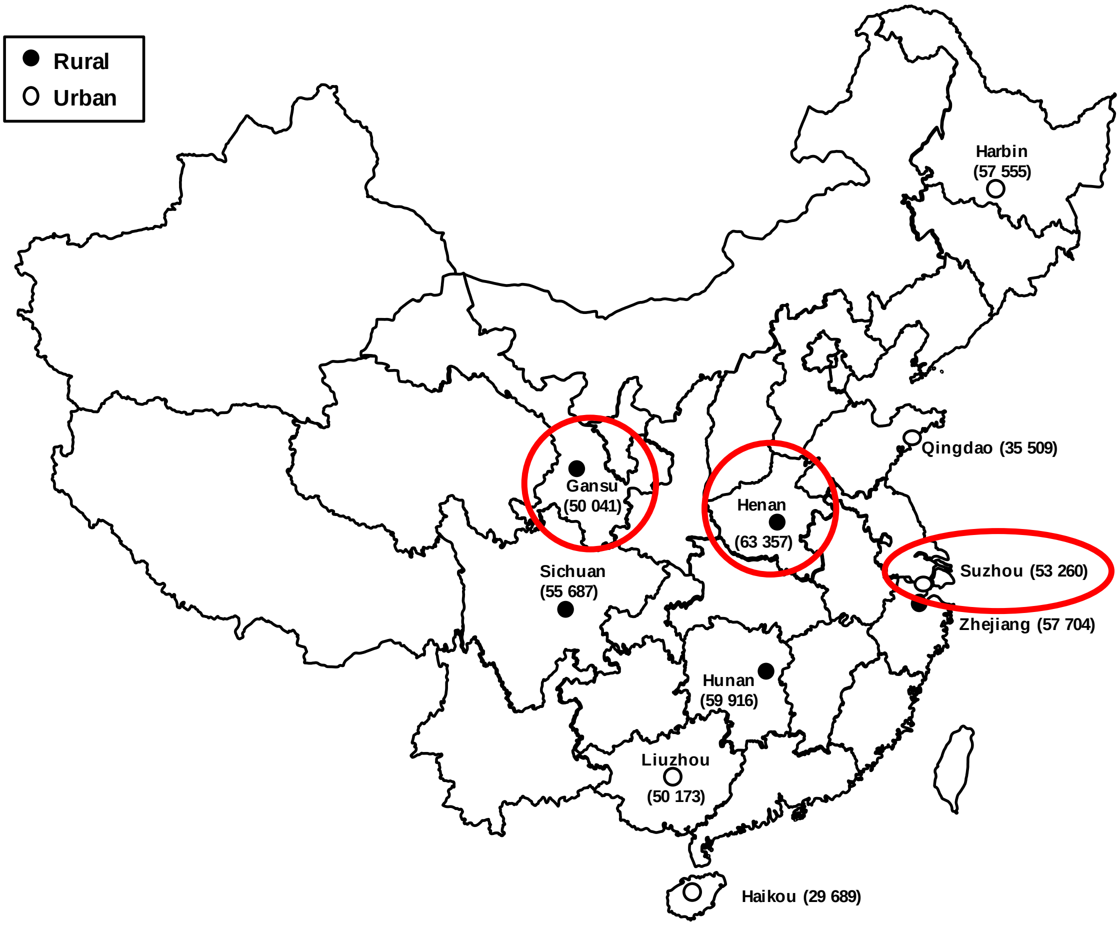

*Figure reproduced from Chen et al. 2011.[1] The figure illustrates the location of the ten study areas of the China Kadoorie Biobank Cohort Study, with black circles indicating the rural sites, and open circles indicating the urban sites. Numbers in brackets are the baseline sample size per study site. The one urban (Suzhou) and two rural (Gansu, Henan) sites included in the CKB-Air study are highlighted in red. This figure is for illustrative purpose and does not represent the exact geography of the wider region.

**eFigure 2. Flowchart of PATS data exclusion by season and device location**

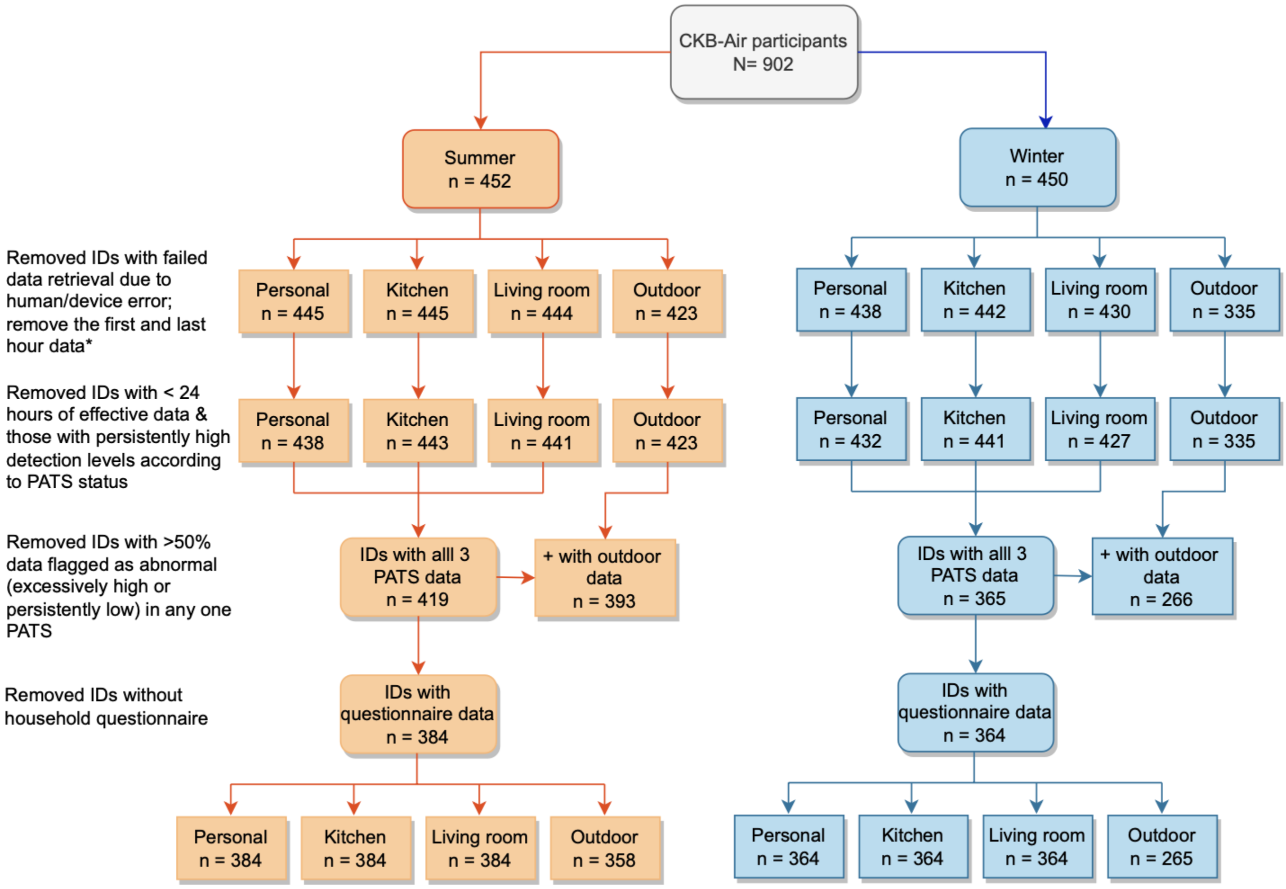

*To remove data likely to be influenced by the initial device deployment and final device collection work.

**eFigure3. Age- and sex-adjusted geometric mean $PM_{2.5}$ concentrations (μg/$m^3$) recorded in the personal, kitchen, living room, and community monitors by season, primary cooking fuels**

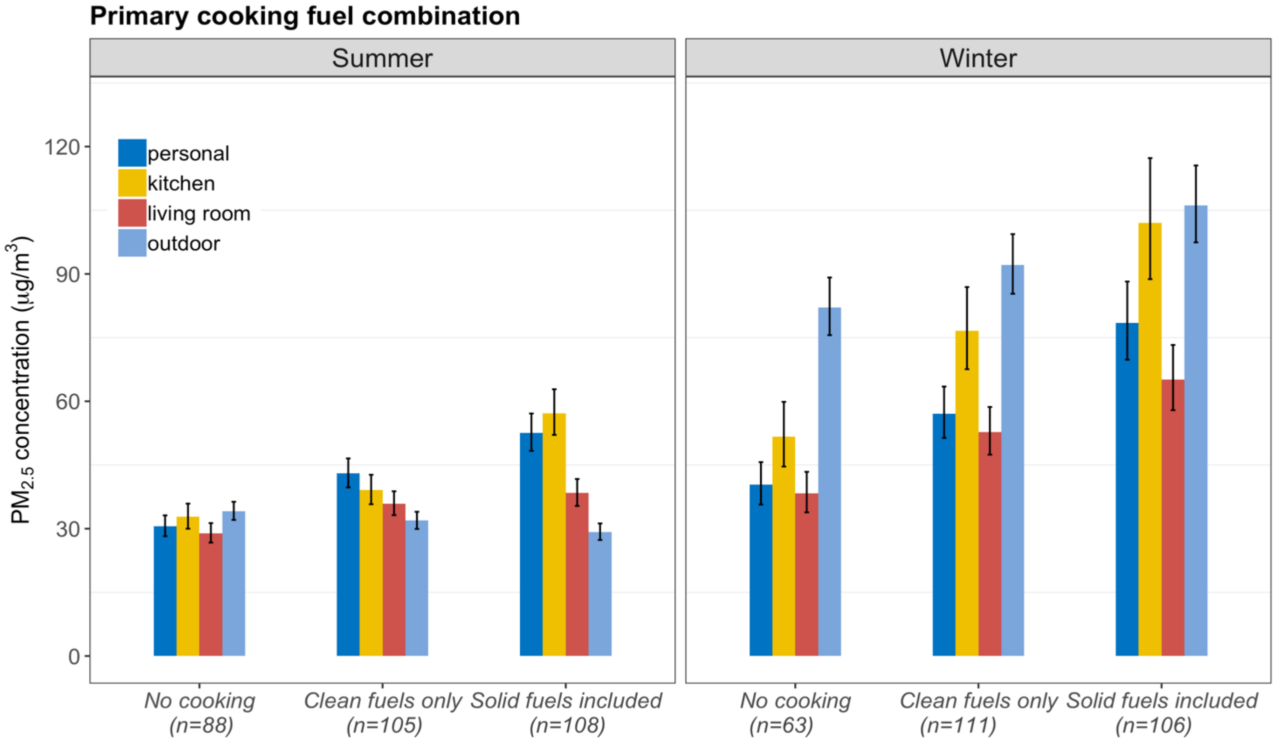

Note: The subjects were restricted to those frequently cooking only; "No cooking" refers to those who reported that no fuels were used for cooking. Each vertical bar represents adjusted geometric means of each location by exposure groups, with vertical black lines showing the corresponding 95% confidence intervals (CIs). Non-overlapping CIs between bars indicate statistically significant difference. From left to right the four bars in each group are personal, kitchen, living room, and community $PM_{2.5}$ levels.

**eFigure 4A. The correlation matrix between the concentrations of $PM_{2.5}$ for personal, kitchen, living room and community environments by cooking & heating fuel combination with no fuel used for cooking or heating**

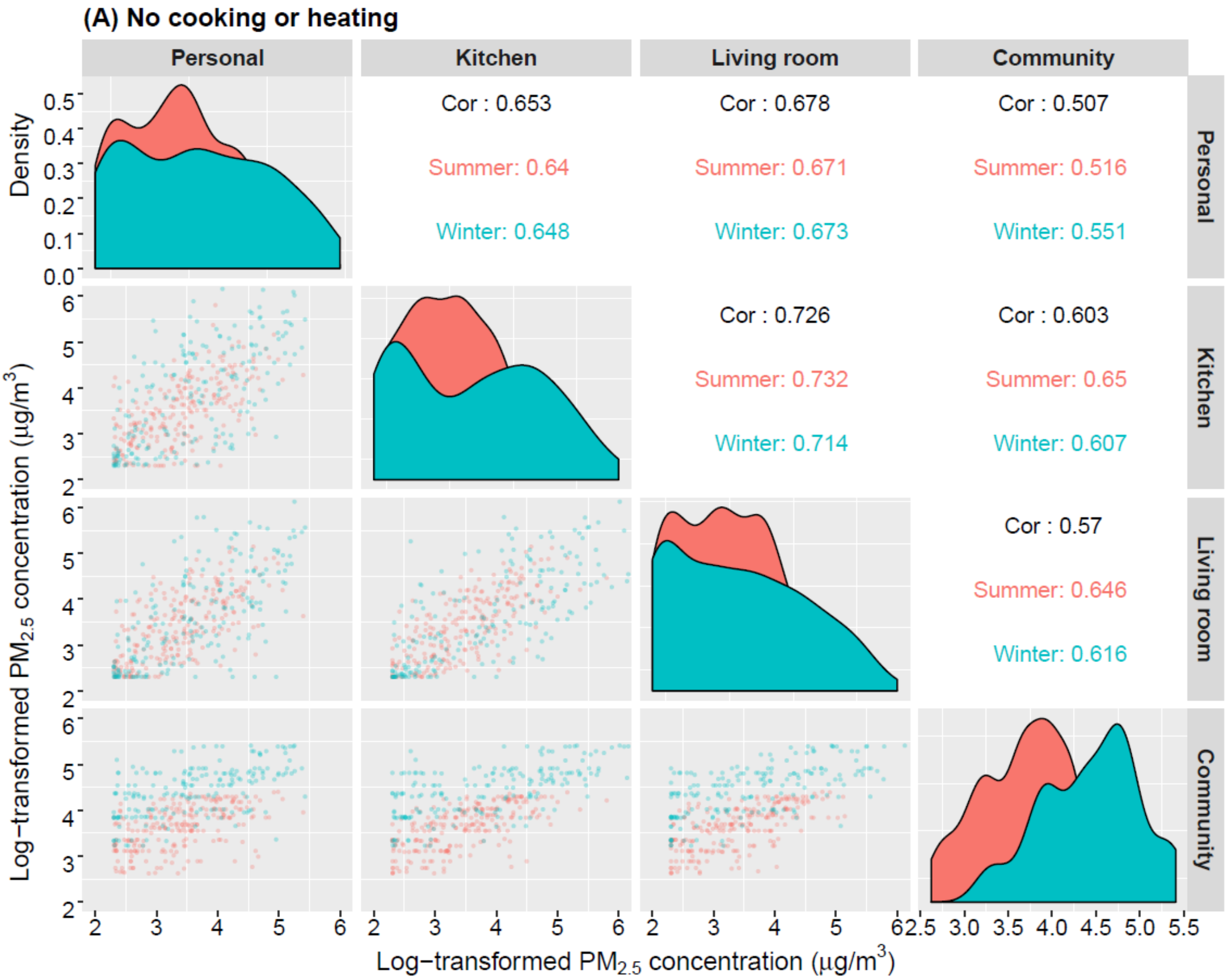

Note: Red area under curves and dots are summer data; blue area under curves and dots are winter data; black numbers in boxes are overall Spearman correlation coefficient; red and blue numbers are summer- and winter-specific correlation.

**eFigure 4B. The correlation matrix between the concentrations of $PM_{2.5}$ for personal, kitchen, living room and community enivornments by cooking & heating fuel combination with clean fuels**

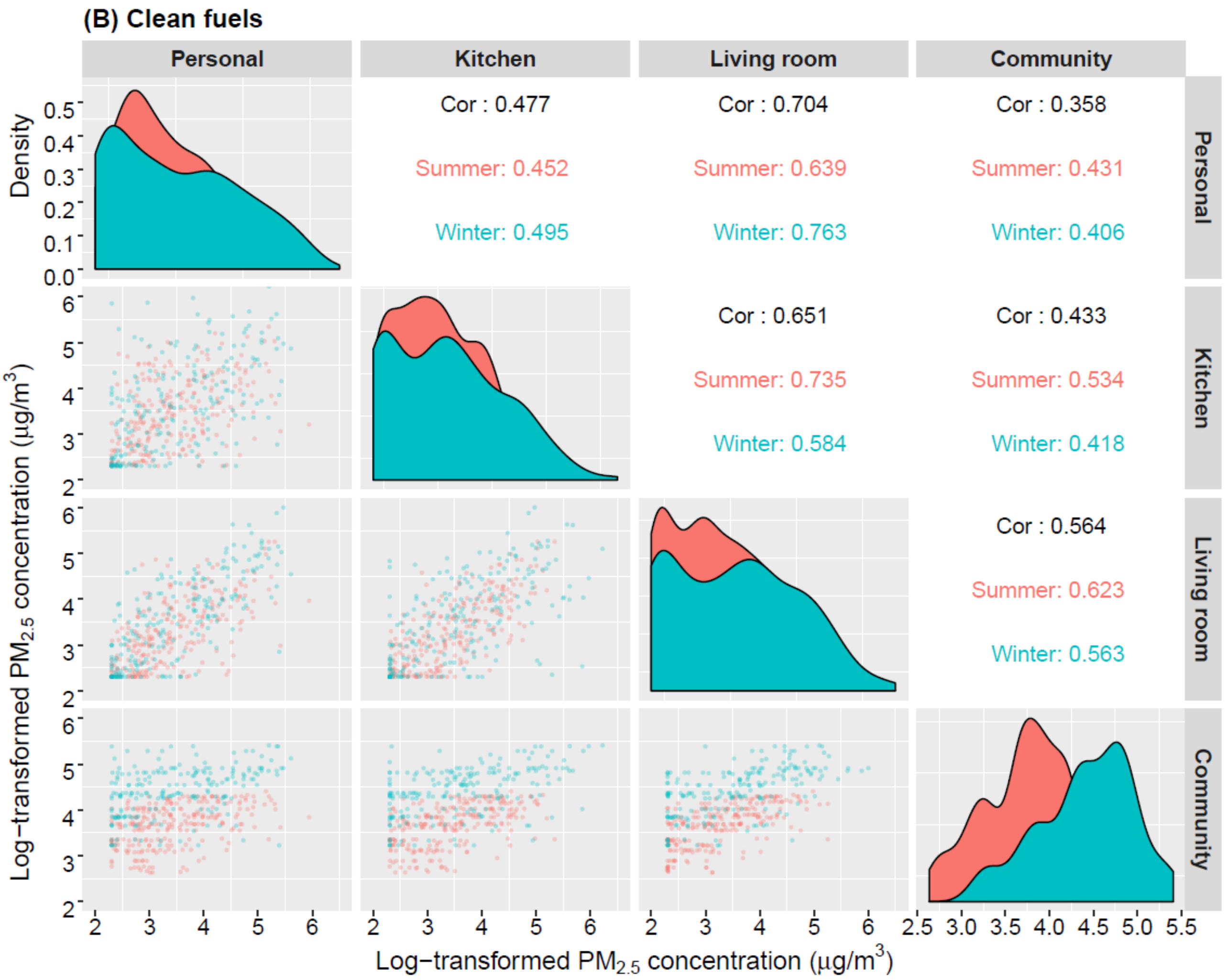

Note: Red area under curves and dots are summer data; blue area under curves and dots are winter data; black numbers in boxes are overall Spearman correlation coefficient; red and blue numbers are summer- and winter-specific correlation.

**eFigure 4C. The correlation matrix between the concentrations of $PM_{2.5}$ for personal, kitchen, living room and community environments by cooking & heating fuel combination with solid fuels included**

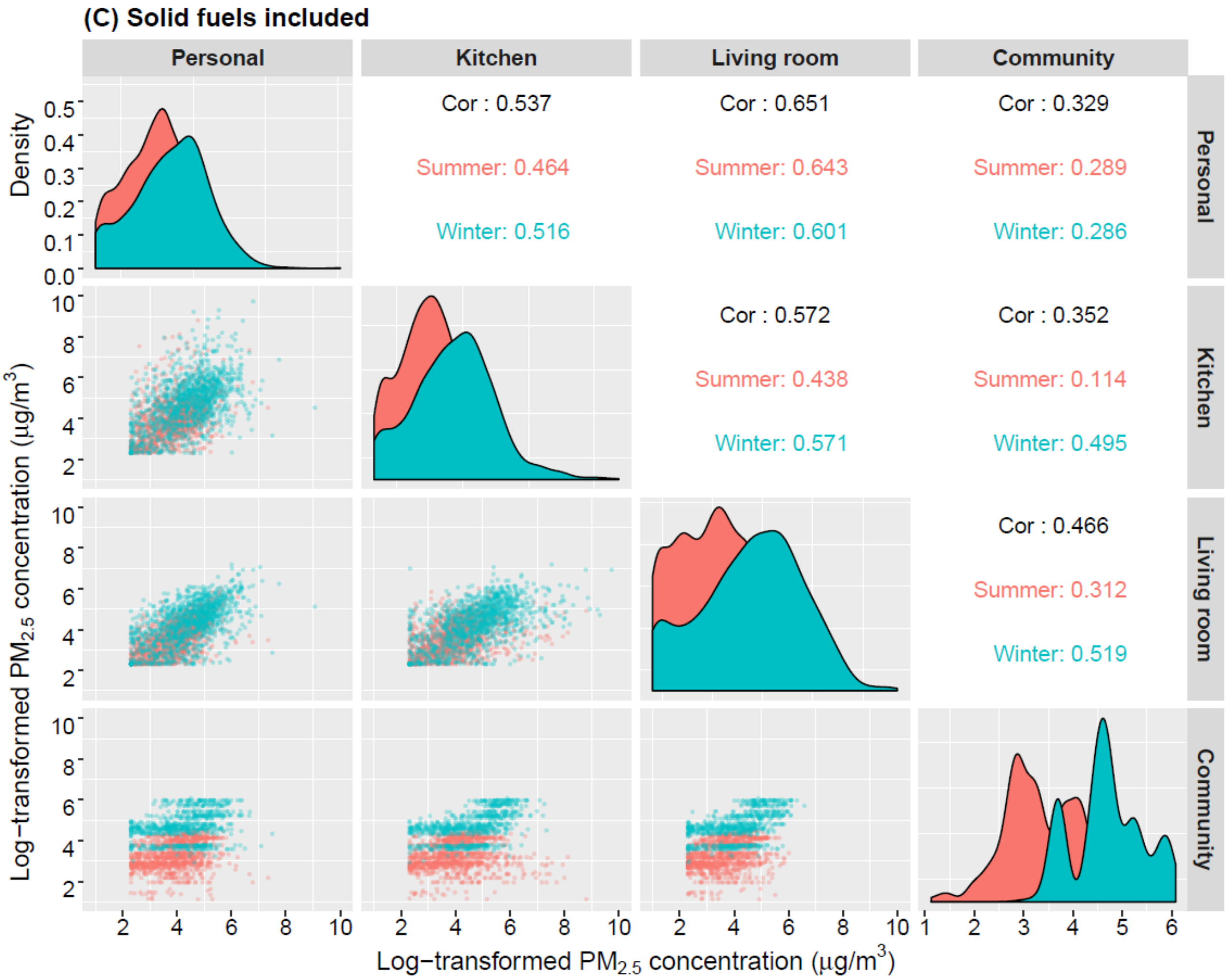

Note: Red area under curves and dots are summer data; blue area under curves and dots are winter data; black numbers in boxes are overall Spearman correlation coefficient; red and blue numbers are summer- and winter-specific correlation.
